# The Infection Rate of the Coronavirus Disease 2019 (COVID-19) in Wuhan, China

**DOI:** 10.1101/2020.05.02.20088724

**Authors:** Hui-Qi Qu, Zhangkai J. Cheng, Zhifeng Duan, Lifeng Tian, Hakon Hakonarson

## Abstract

**What is already known about this subject?:** The Wuhan city in China had a much higher mortality rate (Feb 10^th^ statistics: 748 death/18,454 diagnosis =4.05%; Apr 24^th^ statistics: 3,869 death/50,333 diagnosis=7.69%) than the rest of China.

**What are the new findings?:** Based on our analysis, the number of infected people in Wuhan is estimated to be 143,000 (88,000 to 242,000) in late January and early February, significantly higher than the published number of diagnosed cases.

**What are the recommendations for policy and practice?:** Increased awareness of the original infection rates in Wuhan, China is critically important for proper public health measures at all levels, as well as to eliminate panic caused by overestimated mortality rate that may bias health policy actions by the authorities

---

The pandemic of the Coronavirus Disease 2019 (COVID-19) started in Wuhan, China in December 2019. Wuhan is a megacity with a population of about 11 millions. To prevent the spread of this alarming infectious disease, the whole city was locked down by the government since Jan 23^rd^, 2020. However, despite the efforts, COVID-19 has spread to many countries across the world, and is causing an unprecedented pandemic as well as serious public health concern considering its high mortality rate. According to the large-sample analysis by Wu and McGoogan(1, 2), the case-fatality rate (CFR) was 2.3% in China, i.e. 1023 of 44-672 confirmed cases by February 11, 2020 with a significant proportion of the cases from Wuhan. The large number of infected people in the city of Wuhan had put a tight strain on essential medical resources. The city had a much higher mortality rate (Feb 10^th^ statistics: 748 death/18,454 diagnosis =4.05%; Apr 24^th^ statistics: 3,869 death/50,333 diagnosis=7.69%) than the rest of China. Therefore, the overall CFR of 2.3% in China was likely overestimated, due to strained medical resources and a large number of undiagnosed patients. According to the most recent study, 78% infections were asymptomatic(3). Therefore, a large number of asymptomatic infections in Wuhan might have never been diagnosed, which contributed to the overestimated CFR. To get an accurate estimation of infections is therefore important to assess the CFR in Wuhan precisely.

Using Markov Chain Monte Carlo methods, Wu et al. estimated that 75,815 individuals (95% CI: 37,304, 130,330) had been infected in Wuhan as of Jan 25, 2020(4). After this study, a number of foreign governments evacuated their citizens and performed thorough etiological tests for these people. The evacuation of people can serve as a “random” sample to estimate the infection rate in Wuhan. Using these publicly available data (Table 1), we performed a combined analysis of the infection rates of these population samples, instead of a simple pooled calculation, considering potential different life styles and pathogen exposures across different populations. The combined analysis was done using the Comprehensive Meta-Analysis Software (CMA).

**Table 1.** Number of infected people from different countries.

| Country | Evacuation date | Confirmed cases | Evacuated number | Data sources |
| --- | --- | --- | --- | --- |
| Japan | NA | 9 | 566 | https://www.mhlw.go.jp/stf/newpage_09396.html |
| Korea | 31-Jan | 1 | 368 | http://world.kbs.co.kr/service/news_view.htm?lang=c&Seq_Code=66621<br>https://www.mohw.go.kr/eng/nw/nw0101vw.jsp?PAR_MENU_ID=1007&MENU_ID=100701&page=1&CONT_SEQ=352718<br>http://www.mohw.go.kr/react/al/sal0301vw.jsp?PAR_MENU_ID=04&MENU_ID=0403&page=3&CONT_SEQ=352645 |
| Germany | 1-Feb | 2 | 124 | https://www.dw.com/en/coronavirus-german-evacuation-flight-from-china-carried-two-infected-people/a-52229955 |
| Singapore | 30-Jan | 1 | 92 | https://www.nst.com.my/world/world/2020/01/561086/coronavirus-countries-evacuate-citizens-china<br>https://www.straitstimes.com/singapore/some-singaporeans-with-symptoms-of-virus-staying-behind-in-wuhan-even-as-92-are-evacuated<br>https://www.straitstimes.com/singapore/coronavirus-2-new-cases-in-singapore-including-certis-cisco-staff-who-had-served |
| Italy | 2-Feb | 1 | 56 | https://www.aa.com.tr/en/europe/italy-reports-third-confirmed-case-of-coronavirus/1726934 |
| US | 29-Jan | 0 | 195 | https://edition.cnn.com/2020/02/10/us/coronavirus-american-evacuees-release/index.html |
| Total | NA | 14 | 1401 |  |

Shown in our analysis, there is no significant heterogeneity across different population samples (heterogeneity test P=0.491). The combined infection rate (95% CI)=0.013 (0.008, 0.022) (Fig.1). Based on these results, the number of infected people in Wuhan is estimated to be 143,000 (88,000 to 242,000), or significantly higher than the estimate by Wu et al(4). By our estimation, it is safe to say that a large number of infections in Wuhan, China have not been diagnosed (the number of undiagnosed in late January and early February is much more than the final diagnosed number 50,333 to date), causing the CFR being over-estimated. In addition, our estimation suggests that the lower CFR (0.51%) estimated by the Centre for Evidence-Based Medicine(2) doesn’t mean viral variants and loss of virulence. Taken together, increased awareness of the original infection rates in Wuhan, China is critically important for proper public health measures at all levels, as well as to eliminate panic caused by overestimated mortality rate that may bias health policy actions by the authorities.

**Figure 1.**
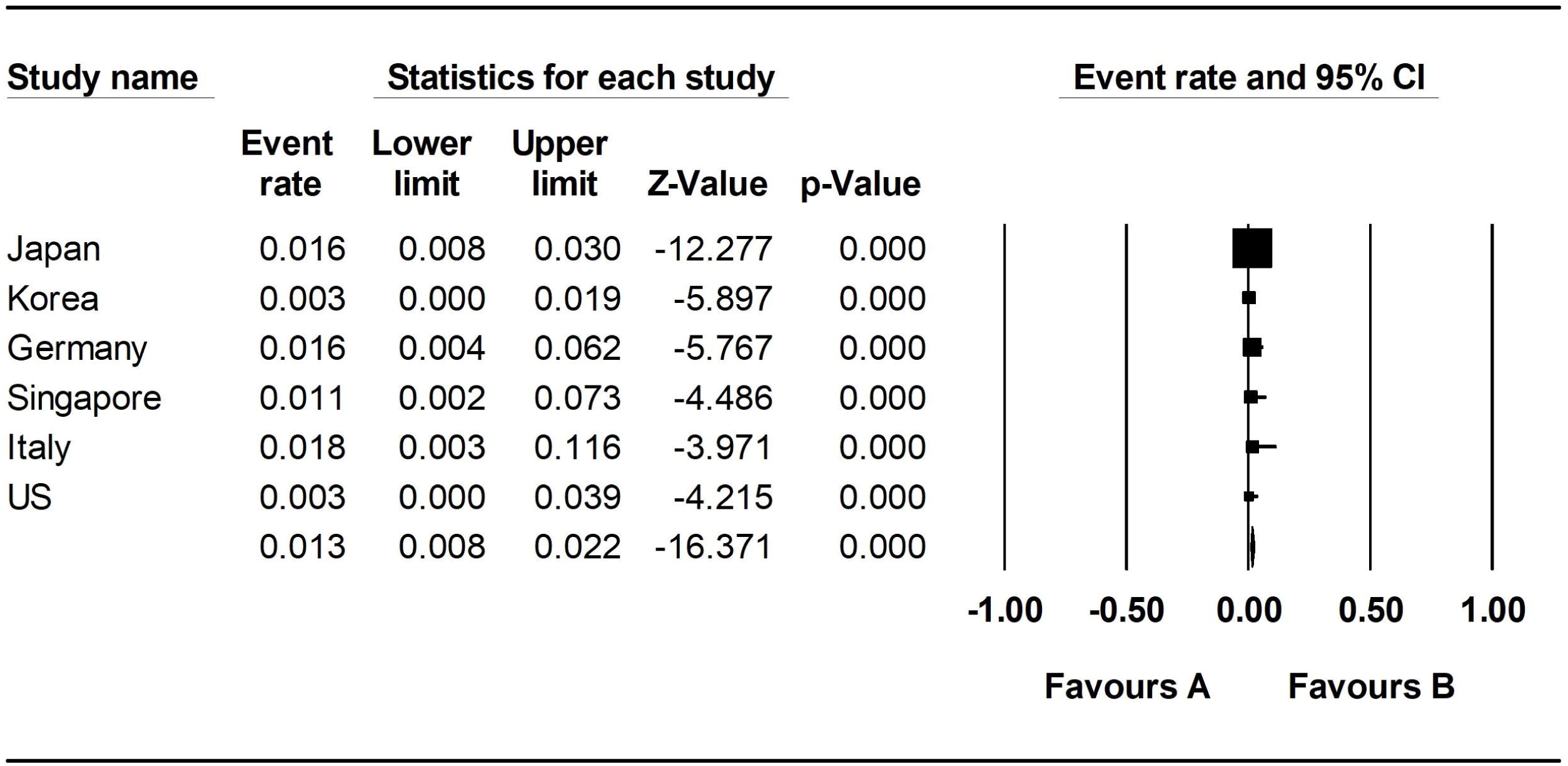
Combined analysis of infection rates of different populations

## Data Availability

Data sharing statement: no additional data.

## Contributorship

H.Q.: literature search, study design, data collection, data analysis, data interpretation, writing

Z.J.C:data collection

Z.D.:data interpretation

L.T.:study design, data interpretation

H.H.:study design, data interpretation, writing

## Data sharing statement

no additional data.

